# Eye care services and Factors associated with levels of satisfaction among patients attending eye clinics at kcmc 2024

**DOI:** 10.1101/2025.09.22.25336383

**Authors:** Newton Yongolo, Furahini Godfrey Mndeme, Jovin R Tibenderana, Andrew Makupa, Sarah Kweka

## Abstract

**Background:** In a hospital setting, patient satisfaction with medical services is a key determinant of clinical efficacy and service quality. There is little data on patient satisfaction with ophthalmic services in Tanzania, which emphasizes the need for focused enhancements in the provision of eye care.

**Objective:** To evaluate the levels of satisfaction with eye care services and the factors associated with satisfaction among individuals attending outpatient clinics at KCMC from August 2023 to July 2024.

**Methodology:** This was a hospital-based analytical cross-sectional study, conducted at KCMC hospital involving 385 individuals from August 2023 to July 2024.The outcome was satisfaction with eye services. A binary logistic regression model was employed to identify independent significant factors influencing satisfaction, with corresponding odds ratios (OR) and significance was declared at p-value of < 0.05. Data were managed and analysed using SPSS 24

**Results:** Overall, patient satisfaction with eye care services was high at 91%. Among specific eye conditions, diabetic retinopathy and conjunctivitis had the highest satisfaction rates of services provided at 94% and 92%, respectively. Staff dressings (OR:7.19; 95% CI: 3.04 - 17.05), communication (OR: 2.47; 95% CI: 1.36-4.49), consultation time (OR: 4.47; 95% CI: 2.05-9.71), patient confidentiality (OR: 2.79; 95% CI:1.50-5.17), affordability of services (OR:2.15; 95% CI: 1.08-4.24), and staff attitudes(OR: 2.64; 95% CI:1.28-5.44) were all independent significant factors associated with satisfaction of eye services.

**Conclusion:** The study found high patient satisfaction with eye care services at KCMC, particularly for diabetic retinopathy, conjunctivitis, and glaucoma, driven by clinical quality and strong interpersonal factors such as professional staff, clear communication, adequate consultation time, confidentiality, affordability and attitudes highlight the importance of both clinical competence and interpersonal aspects in enhancing patient experiences within ophthalmic care.

## Background

It is commonly acknowledged that one of the most important ways to learn about patients’ opinions of the medical care they receive is to measure their level of satisfaction. It is now used all over the world as a measure of service quality, high quality healthcare should align with patient expectations, leading to improved treatment adherence, responsible resource utilization, and faster recovery [1]. The degree to which a patient is content with their healthcare provider is measured by patient satisfaction[2].

To improve the standard of care given, the causes of the discontent must be identified and removed, patient satisfaction surveys have been used in the healthcare delivery industry to accomplish three main objectives. First, they evaluate the impact of the patient’s satisfaction with the services received on their decision-making. Second, it has been thought that improving patient satisfaction plays a significant role in raising the standard of healthcare results. Finally, the surveys help to improve the services provided, increase liability, and comprehend the needs of the patient [2,3].

Eye conditions have grown to be a major public health issue in recent years[4]. Globally, blindness or visual impairment affects at least 2.2 billion people worldwide. Approximately 1 billion of these cases could have been treated or prevented, an estimated 253 million people have moderate to severe visual impairment, and 36 million are totally blind [5]. According to estimates, over 80% of people in sub-Saharan Africa suffer from untreated near vision impairment[6].

In Tanzania, a considerable proportion of the population suffers from blindness and vision impairment; an estimated 290,000 people are blind, and 8.2 million have some kind of vision loss. Even though 80 percent of blindness is thought to be preventable or treatable[7]. Although KCMC plays a vital role in providing ophthalmic care, it is still unknown how satisfied people are with these services, little is known about KCMC’s ophthalmology clinic patients’ satisfaction levels, particularly with regard to specialized eye care services. Therefore, this study aimed to evaluate the levels of satisfaction with eye care services and the factors associated with patients’ satisfaction among individuals attending outpatient clinics at KCMC from August 2023 to July 2024.

## Methodology

### Study design

This was a hospital-based analytical cross-sectional study conducted between 13th August 2023 to 30th July 2024.

### Study setting

KCMC serves a population of about 15 million people, including local women and referred cases from neighboring regions such as Arusha, Tanga, and Manyara. Moshi Municipality has a population of 221,172 with a high density of 3,409 persons per km^2^, while the Kilimanjaro region has a population of 1,861,934 with a density of 124 people per km^2^.The primary socio-economic activities in Kilimanjaro encompass tourism, agriculture and industrial activities [8–10]. The eye department offers pediatric and adult clinics and serves as a training center for MMED in ophthalmology, optometrists, ophthalmic nurses, ophthalmic assistants, and medical students. The adult and pediatric clinics serve about 14000, and 850 patients per year, and the clinics are scheduled throughout the week, with general clinics on Mondays, Tuesdays, and Thursdays, glaucoma and corneal clinics on Tuesdays and Thursdays, and diabetic clinics on Fridays. Surgeries are typically performed from Tuesdays to Fridays and occasionally on Mondays. The pediatric clinic runs throughout the week. The department also offers specialized services including cataracts, glaucoma, retinal, corneal, and strabismus care.

### Study population, sample size and sampling

The study included all patients who attended eye clinics to receive treatment at KCMC Ophthalmology department from August 2023 to July 2024 and convenience sampling was used to recruit the participants. The sample size was calculated by using the maximum satisfaction response of 57% in a study conducted in Ethiopia [11]. The Cochran formula below was used to calculate sample size

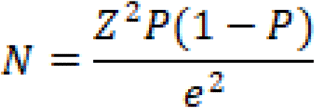

Where

N= Minimum sample size

Z value corresponding to 95% confidence interval (marginal error=1.96).

P= Proportion of satisfaction used was 57%

E= Marginal error (0.05)

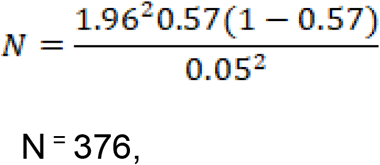

Hence the minimum sample size was 376 patients

### Eligibility criteria

All patients attending outpatient eye clinics, including those presenting for first visits and follow-up appointments, were included in the study; the sole exclusion criterion applied was unwillingness to participate, whereby patients who declined to provide consent were excluded from the analysis

### Variables

The dependent variable for this study was satisfaction of the eye care services which was dichotomized into satisfied and not satisfied. The independent variables included sociodemograpic characteristics (Age, Education level, Occupation, Marital status) and the associated factors (Waiting time, Consultation time, Physical facilities, Dressing code, Accessibility of services, Attitude, Communication, Doctor-patient relationship,Appointments, Costs of services, Explanations of eye services, Privacy, politeness and Eye care services)

### Data processing and analysis

Data cleaning and analysis performed using SPSS Version 24. Exploration of the data was done to check for duplicates, unusual observations, and missing values prior to analysis. Based on previous literatures, some of the variables were generated and some re-categorized for better comparability and clarification. Descriptive statistics were summarized using frequencies and percentages. Mean and standard deviation were used to summarzide continous varibales. Differences in satisfaction levels based on socio-demographic characteristics were assessed using the Chi-square test. A binary logistic regression model was employed to identify independent significant factors influencing satisfaction, with corresponding odds ratios (OR) and 95% confidence intervals (CI) provided. A p-value of less than 0.05 was considered statistically significant.

### Ethical consideration

The study was conducted with ethical clearance obtained from the Kilimanjaro Christian Medical University College Research and Ethics Review Committee (with number PG 90/2023). Data collection was approved by the Director of Hospital Services through the head of the Department of Ophthalmology at KCMC. Before data collection, informed consent was obtained from all participating patients. To ensure confidentiality and data protection, the use of encrypted computer passwords was implemented, with access restricted to the researcher and supervisors only.

## Results

### Social demographic characteristics of the study participants

A total of 385 study participants were included in the study. The majority of participants were in the 40-59 age group (24.9%, n=96), with slightly more male participants (51.2%, n=197). The largest portion of the participants were single (44.7%, n=172), and most had a primary level of education (35.1%, n=135). More than half of the participants resided in urban areas (53.2%, n=205), with a significant portion being unemployed (40.3%, n=155). Additionally, over half of the participants (57.9%, n=223) reported waiting for more than one hour for their consultation (**Table 1**).

**Table 1:** Social demographic characteristics of study participants (N=385)

| **Variable** | **Frequency** | **Percentage** |
| --- | --- | --- |
| **Age (Mean±SD)** | 34.6±23.95 |  |
| **Age group (years)** |  |  |
| ≤5 | 59 | 15.3 |
| 6 -15 | 67 | 17.4 |
| 16 – 39 | 87 | 22.6 |
| 40 – 59 | 96 | 24.9 |
| 60+ | 76 | 19.7 |
| **Sex** |  |  |
| Male | 197 | 51.2 |
| Female | 188 | 48.8 |
| **Marital status** |  |  |
| Married | 172 | 44.7 |
| Widowed | 47 | 12.2 |
| Separated | 34 | 8.8 |
| Divorced | 15 | 3.9 |
| Single | 117 | 30.4 |
| **Education level** |  |  |
| No formal | 67 | 17.4 |
| Primary | 135 | 35.1 |
| Secondary | 114 | 29.6 |
| Tertiary | 69 | 17.9 |
| **Residence** |  |  |
| Urban | 205 | 53.2 |
| Rural | 180 | 46.8 |
| **Occupation** |  |  |
| Employed | 73 | 19.0 |
| Self-employed | 118 | 30.6 |
| Unemployed | 155 | 40.3 |
| Retired | 39 | 10.1 |
| **Time taken waiting for consultation** |  |  |
| Less than 30 min | 24 | 6.2 |
| 30 min – 1 hr | 138 | 35.8 |
| More than 1 hr | 223 | 57.9 |
| **Estimated consultation time (min)** |  |  |
| 10 – 15 | 158 | 41.0 |
| 16 – 20 | 114 | 29.6 |
| More than 20 | 113 | 29.4 |
| **Distribution of eye condition** |  |  |
| Glaucoma | 83 | 21.6 |
| Cataract | 51 | 13.2 |
| Conjunctivitis | 80 | 20.8 |
| Diabetes | 39 | 10.1 |
| Others | 132 | 34.3 |

### Level of Satisfaction of Eye Care Services According to five Domains of Patient Satisfaction

In the empathy domain, the total mean score was 15.67 (±2.76), while in the responsiveness domain, it was 13.47 (±2.66). The satisfaction rates for eye care services in various domains were as follows: Tangibility - 92.5%, Reliability - 81.8%, Responsiveness - 79.7%, Assurance - 83.9%, and Empathy - 66.5% (**Table 2**).

**Table 2:** Level of satisfaction with eye care services according to five domains of patient satisfaction.

| Domain | Domain cutting points | Mean (SD) | Min-Max Domain Scores | 95%CI | Level of satisfaction |  |
| --- | --- | --- | --- | --- | --- | --- |
|  |  |  |  |  | Satisfied | Not satisfied |
| **General satisfaction** | 61.6 | 60.95±7.88 | 22 - 88 | | 351(91.2) | 34(8.8) |
| **Tangible** | 11.2 | 11.98±2.269 | 4 - 16 | 2.029 - 2.568 | 356(92.5) | 29(7.5) |
| **Reliability** | 14 | 11.75±2.26 | 5 - 20 | 1.968 - 2.510 | 315(81.8) | 70(18.2) |
| **Responsibility** | 14 | 13.47±2.66 | 4 - 16 | 2.027 - 2.404 | 307(79.7) | 78(20.3) |
| **Guarantee** | 11.2 | 8.08±1.71 | 4 - 16 | 2.33 - 3.109 | 323(83.9) | 62(16.1) |
| **Empathy** | 16.8 | 15.67±2.76 | 6 - 24 | 1.769 - 2.180 | 256(66.5) | 129(33.4) |

### Overall levels of satisfaction with eye care services among patients in the Ophthalmology clinic

The overall mean score for general satisfaction was 60.95, with a standard deviation of 7.88. Out of 385 patients who visited the eye clinics, 351 (91.2%) reported being satisfied with the eye care services provided, while 34 (8.8%) expressed dissatisfaction. In the tangible domain, the overall level of satisfaction was 356 (92.5%), with 86.2% of patients satisfied with the staff’s personalities and 71.4% content with the accessibility of the eye care services. In the reliability domain, the overall satisfaction level was 315 (81.8%), with 75.3% of patients pleased with the doctors’ good attitude and 73.5% satisfied with the positive doctor-patient relationship. Regarding the responsiveness domain, the overall satisfaction was 307 (79.7%), with 68.8% of patients satisfied with the pediatric specialized services and 67.5% content with the consultations provided at the clinic. In the assurance domain, the overall satisfaction was 328 (83.9%), with 71.7% of patients satisfied with the information provided by the doctors (**Table 3**).

**Table 3:** Satisfaction of patients in different domains of eye care services satisfaction (N=385)

| Satisfaction of item domain | Level of satisfaction | | Mean (±SD) |
| --- | --- | --- | --- |
|  | Satisfied<br>(Strong agree+Agree)<br>n(%) | Not satisfied<br>(Strong disagree+disagree)<br>n(%) |  |
| **Level of satisfaction** | **351(91.2)** | **34(8.8)** | **60.95±7.88** |
| **Tangible** | **356(92.5)** | **29(7.5)** | **11.98±2.26** |
| There are enough doctors at the clinic | 299(77.7) | 86(22.3) | 3.28±1.00 |
| Good physical facilities at the clinic | 223(57.9) | 162(42.1) | 2.78±0.84 |
| Staff dressing is perfectly | 322(86.2) | 63(13.8) | 3.12±0.73 |
| Easy access to eye care services | 275(71.4) | 110(28.6) | 2.79±1.08 |
| **Reliability** | **315(81.8)** | **70(18.2)** | **11.75±2.26** |
| Doctors have good attitudes to patients at the clinic | 290(75.3) | 95(24.7) | 2.97±0.90 |
| Good communication between doctors and patient | 268(69.6) | 117(30.4) | 2.95±0.92 |
| Good relationship between doctors and patients | 283(73.5) | 102(26.5) | 3.05±0.92 |
| Clinic waiting time for services | 252(65.5) | 133(34.5) | 2.78±0.98 |
| **Responsiveness** | **307(79.7)** | **78(20.3)** | **13.47±2.66** |
| The correct time of service provision at the clinic | 191(49.6) | 194(50.4) | 2.43±1.07 |
| Doctors' appointment satisfaction at the clinic | 224(58.2) | 161(41.8) | 2.66±0.99 |
| Doctors' consultation satisfaction at the clinic | 260(67.5) | 125(32.5) | 2.83±0.92 |
| Easy delivery of results of investigations at the clinic | 226(58.7) | 159(41.3) | 2.69±1.02 |
| Satisfaction with pediatric specialized services | 265(68.8) | 120(31.2) | 2.87±0.93 |
| **Guarantee** | **323(83.9)** | **62(16.1)** | **8.08±1.71** |
| Doctors provides good information to the patient | 276(71.7) | 109(28.3) | 2.85±0.98 |
| Eye care services are less expensive | 161(41.8) | 224(58.2) | 2.36±1.01 |
| Doctors provides treatment and drug explanations to patients | 262(68.1) | 123(31.9) | 2.88±1.00 |
| **Empathy** | **256(66.5)** | **129(33.5)** | **15.67±2.76** |
| Presence of confidentiality of doctors and patients at the clinic | 220(57.1) | 165(42.9) | 2.58±1.01 |
| Staff receive money from the patients | 130(33.8) | 255(66.2) | 2.12±1.05 |
| Staff express politeness to the patients | 226(58.7) | 159(41.3) | 2.61±1.00 |
| Doctors pay attention to the patients at the clinic | 262(68.1) | 123(31.9) | 2.81±0.96 |
| Enough time for the patient to meet with Doctors at the clinic | 239(62.1) | 146(37.9) | 2.70±0.972 |
| Satisfied with specialized retina services | 282(73.2) | 103(26.8) | 2.85±0.84 |

### Levels of satisfaction associated with eye care services

Among the 385 patients, 37 (94.9%) of those with diabetic retinopathy were satisfied with the treatment, 74 (92.5%) of those with conjunctivitis were satisfied with the treatment, 124 (93.9%) of patients with other eye diseases were satisfied, and 76 (91.6%) of patients with glaucoma expressed satisfaction with the treatment (**Figure 1**).

**Figure 1:**
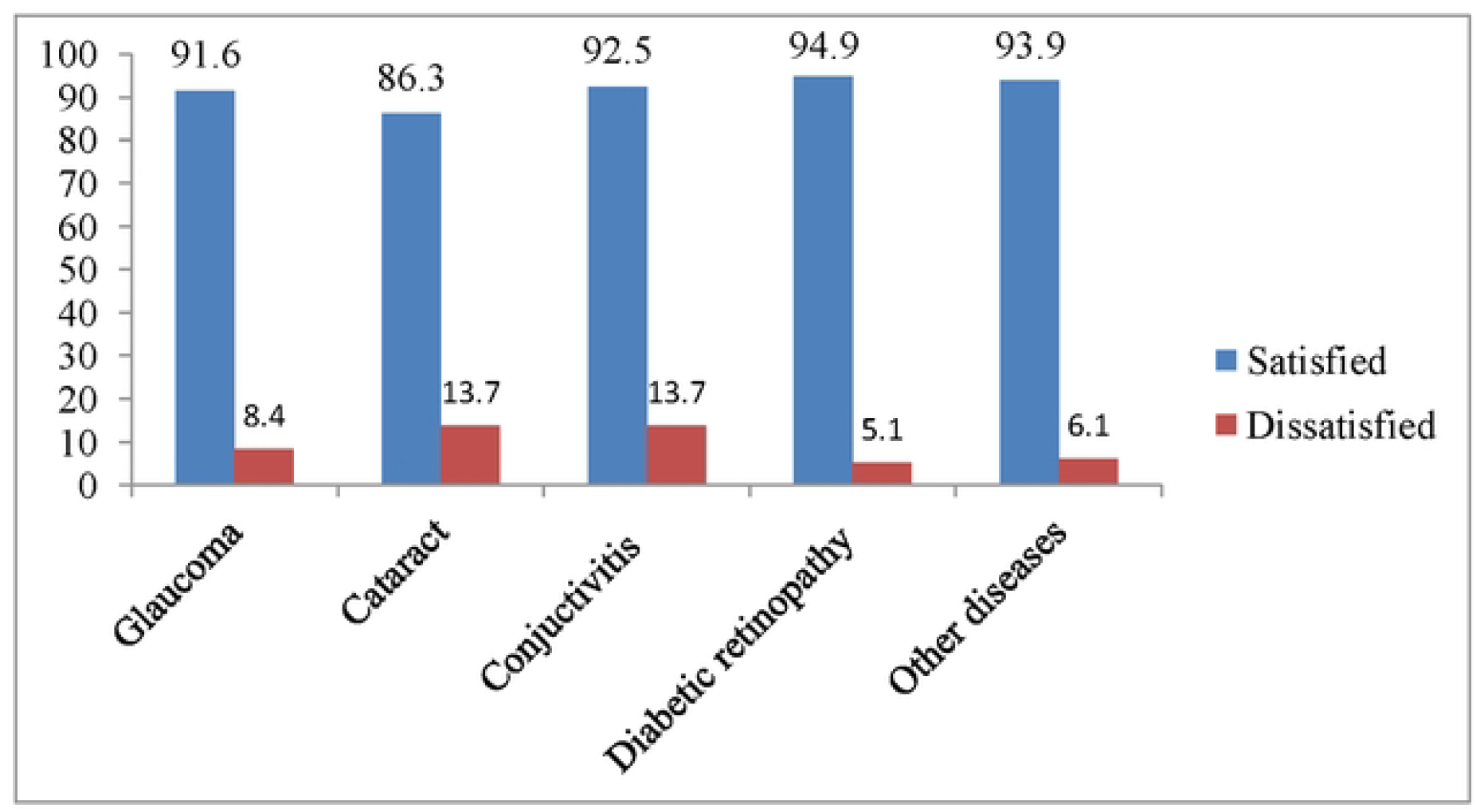
Levels of satisfaction with specialized eye care services (N=385)

### Factors associated with the level of satisfaction among patients attending ophthalmology clinic

The study revealed that staff dressings were associated with a 7.2 times higher likelihood of patient satisfaction with the eye care services at the clinic (AOR= 7.19, 95% CI: 3.04-17.05). Additionally, satisfaction was 2.6 times more likely when staff exhibited a positive attitude towards patients (AOR = 2.64, 95% CI:1.28-5.44). Doctors’ effective communication with patients was linked to a 2.5 times higher likelihood of satisfaction with the clinic’s eye care services (AOR = 2.47, 95% CI: 1.36-4.49). Moreover, the time doctors spent consulting with patients was associated with a 4.5 times higher chance of satisfaction with the services provided at the clinic (AOR = 4.47, 95% CI:2.05-9.71). Patient satisfaction was 2.8 times more likely when there was a high level of confidentiality regarding patient information at the clinic (AOR = 2.79, 95% CI: 1.50-5.17) (**Table 4**).

**Table 4:** Association of the level of satisfaction with eye care services according to the items in the domain.

| **Variables** | **AOR(95%CI)** | **P-value** |
| --- | --- | --- |
| Physical facilities | 1.61(0.68-3.81) | 0.27 |
| Personality | 7.19(3.04-17.05) | <0.001 |
| Staffs attitude | 2.64(1.28-5.44) | 0.008 |
| Communications | 2.47(1.36-4.49) | 0.003 |
| Doctors–patient relationship | 2.25(1.25-4.06) | 0.006 |
| Patients appointment | 1.21(0.70-2.07) | 0.48 |
| Consultation time | 4.47(2.05-9.71) | <0.001 |
| Costs of eye care services | 2.15(1.08-4.24) | 0.02 |
| Explanations of eye services | 1.35(0.76-2.40) | 0.29 |
| Privacy | 2.79(1.50-5.170) | 0.001 |
| Politeness | 3.41(1.65-7.05) | 0.001 |
| Paying attention to patients | 2.57(1.37-4.81) | 0.003 |

## Discussion

The study aimed to evaluate the levels of satisfaction with eye care services and the factors associated with satisfaction among individuals attending outpatient clinics at KCMC from August 2023 to July 2024.

The study reported a high overall patient satisfaction rate of 91% with eye care services. The overall mean score for general satisfaction was 60.95. Almost all (94.9%) of those with diabetic retinopathy were satisfied with the treatment. Factors contributing to this high satisfaction included accessible and affordable services, sufficient staffing, professional appearance, strong doctor-patient relationships, and effective communication.

The high satisfaction rate with eye care services in this study may be attributed to quality care, effective communication, accessibility, affordability, well-equipped facilities and culturally sensitive service delivery. The findings align aligns closely with findings from similar studies in Mozambique (98.3%) and Northern Bangladesh (95%) [12,13]. It is also comparable to a study in southwest Ethiopia, which reported a 97.8% satisfaction rate [14]. These similarities could be explained by the use of comparable data collection tools and methods such as self-administered questionnaires, especially in the Mozambique study and by shared socio-economic conditions and healthcare infrastructure in the case of Ethiopia. Contradicting findings were reported in Nepal, India and Gaza where they found the low satisfaction to the eye services ie 75%, 76% and 63.9% respectively [15–17]. These discrepancies may be due to varying sample sizes such as Nepal’s study with only 82 participants compared to 385 in this study and contextual factors, including political instability in Gaza affecting patient satisfaction, and limited specialist availability and high service costs in Southern India contributing to lower satisfaction levels.

The high satisfaction rates among diabetic retinopathy patients may be attributed to the availability of specialized care provided by skilled and knowledgeable specialists, affordable service costs, and the presence of sufficient residents and support staff at the clinics. The satisfaction rate for diabetic retinopathy treatment in our study was similar to findings by Wirostko [18] and Shayan [19], who reported 83% and 85% satisfaction, respectively, with only slight differences. These variations may be due to differences in data collection methods, as Shayan et al. used telephone interviews, whereas this study used in-person surveys. Additionally, Wirostko et al.’s study involved three Canadian eye centers with different healthcare systems, which could explain the slight variation in satisfaction compared to our study, which focused on a single eye center

The staff dress code showed the highest association with and this can be explained by the fact that a neat and consistent dress code is often seen as a mark of good hygiene and infection control something especially important in clinical settings where close contact is common, like during eye exams or treatments, wearing professional attire also helps build trust and shows patients they’re in capable hands, making them feel more at ease with the care they’re receiving [20], clear guidelines on what staff should wear make it easier to recognize different roles, which reduces confusion and creates a more organized, professional atmosphere. For many patients especially older adults or those who aren’t familiar with medical environments this kind of visual consistency can provide real comfort and help ease anxiety.

A warm and positive attitude from staff plays a huge role in how satisfied patients feel with their eye care[21]. When healthcare providers are friendly, respectful, and show empathy, it makes patients feel seen, heard, and cared for especially important when someone is worried about losing their vision or dealing with an unfamiliar condition[22]. Taking the time to listen, explain things clearly, and answer questions helps patients feel more involved in their own care, which can make a big difference in their overall experience. Even in busy clinics or places with limited resources, kindness and compassion go a long way in making patients feel comfortable and supported. The findings of this study are consistent with the studies done in Nigeria and Iran [23,24] where they highlighted the importance of respect, empathy, and supportive behavior in promoting patient satisfaction.

Consultation duration was a significant factor, echoing findings from Ethiopia [14] and Saudi Arabia [25] suggesting that longer consultations are associated with better communication, detailed examination, and increased patient trust. The possible reason for these findings could be longer consultation time allows more time for better clinical assessments, clearer explanations, and more personalized attention. In eye care, where precise diagnoses and effective treatments rely heavily on detailed exams and understanding a patient’s history, taking the time truly makes a difference in both the quality of care and how well patients understand their condition[26]. Longer visits also give patients the chance to ask questions, share their worries, and feel like they’re being listened to something that builds trust and strengthens the relationship between patient and provider. When consultations aren’t rushed, patients are more likely to feel that their care is thoughtful and respectful, which plays a big role in how satisfied they are with the entire experience

As highlighted in this study, the relationship between a doctor and patient plays a crucial role in how satisfied patients feel with their eye care. At its best, this relationship is built on trust, empathy, respect, and clear communication making patients feel at ease and confident in the care they’re receiving[27]. In eye care, where treatment plans can be complex or require long-term follow-up, a strong connection with the doctor helps patients feel more involved and supported, which in turn encourages them to stick with their treatment and feel good about the results. When patients sense that their doctor truly cares, takes the time to listen, and explains things in a way they can understand, it makes them feel respected and valued greatly improving their overall experience and satisfaction with the care they receive[28]. Similar findings were reported from previous study [29] which indicated that the doctor-patient relationship significantly predicted satisfaction levels for outpatients.

## Conclusion and Recommendation

Overall satisfaction was high (91.2%), with consistently high levels across various eye conditions. While most socio-demographic factors showed no significant association with satisfaction. Key service-related factors significantly associated with satisfaction included staff dress code (strongest influence), staff attitude, doctor-patient communication, consultation time, confidentiality, affordability, and respectful interaction highlighting the importance of interpersonal and service quality factors in shaping patient satisfaction. Key recommendations from the study include enhancing doctor–patient relationships, maintaining positive staff attitudes, reducing waiting times through staff training, ensuring confidentiality, offering flexible service hours, and keeping services affordable to sustain and further improve patient satisfaction with eye care services at KCMC.

## Acknowledgments

Special appreciation is extended to Dr. William Makupa and Dr. Furahini Mndeme for their invaluable insights, corrections, and contributions that significantly enriched this work. I am also thankful to my colleagues and fellow residents for their encouragement and support during the preparation of this research.

## List of Abbreviation

AOR: Adjusted Odds Ratio
ECF: Eye Care Foundation
CI: Confidence Interval
KCMC: Kilimanjaro Christian Medical Centre
SSA: Sub Saharan Africa
KCMC: Kilimanjaro Christian Medical University College
SD: Standard Deviation
WHO: World Health Organization

## Data availability

The data generated and analysed during the current study are available from the corresponding author upon reasonable request. Due to ethical and confidentiality considerations, the data are not publicly shared.

## Authorship contributions

Newton Yongolo: Conceptualization, Data curation, Formal analysis, Methodology, Visualization, Writing – original draft, Writing – review & editing. Sarah Kweka. Jovin R Tibenderana: Methodology Writing – review & editing. Andrew Makupa: Writing – original draft, Writing – review & editing, Supervision.

